# How many labels do I need? Self-supervised learning strategies for multiple blood parasites classification in microscopy images

**DOI:** 10.1101/2024.02.29.24303535

**Authors:** Roberto Mancebo-Martín, Lin Lin, Elena Dacal, Miguel Luengo-Oroz, David Bermejo-Peláez

## Abstract

Bloodborne parasitic diseases such as malaria, filariasis or chagas pose significant challenges in clinical diagnosis, with microscopy as the primary tool for diagnosis. However, limitations such as time-consuming processes and the dependence on trained microscopists is critical, particularly in resource-constrained settings. Deep learning techniques have shown value to interpret microscopy images using large annotated databases for training. In this work, we propose a methodology leveraging self-supervised learning as a foundational model for blood parasite classification. Using a large unannotated database of blood microscopy images, the model is able to learn important image representations that are subsequently transferred to perform parasite classification of 11 different species of parasites requiring a smaller amount of labeled data. Our results show enhanced performance over fully supervised approaches, with ∼100 labels per class sufficient to attain an F1 score of ∼0.8. This approach is promising for advancing in-vitro diagnostic systems in primary healthcare settings.

## 1. Introduction

Infectious diseases caused by blood parasites, including filariae, malaria, and chagas disease, continue to pose significant health challenges worldwide. The accurate and timely identification of these parasites is crucial for effective patient management, disease surveillance, and public health interventions. While microscopy remains the primary tool method for the diagnosis of those diseases, it is hindered by its time-consuming nature and reliance of substantial human experts, which is often limited in resource-constrained settings, and difficult to achieve for the correct identification of those less common parasites[1], [2].

In this context, the integration of artificial intelligence (AI) into the analysis of microscopy images emerges as a transformative solution. AI-driven can help automating the detection and classification of blood parasites, thereby mitigating the challenges associated with manual examination. This paradigm shift not only expedites the diagnostic process but also addresses the shortage of skilled personnel.

Despite the considerable efforts in this field, all previously proposed AI-based systems for parasite detection in microscopy blood images focus on specific parasites without considering the potential coexistence of various parasites within the same sample. The majority of methods exhibit this targeted approach, particularly in the context of malaria [3]–[6], while few methods address diseases like Chagas or leishmania [7]–[9], and none for filariasis. The absence of a universal method to detect any blood parasite in a given sample is a significant limitation, particularly in areas with coinfection[10], [11].

Development of AI algorithms for medical images traditionally faces the critical need for abundant labeled data for effective training. In response to this challenge, self-supervised learning (SSL) has emerged as a compelling alternative to conventional supervised methods by learning features from a large, unlabeled dataset can serve as a foundation for downstream tasks like classification, reducing the required number of labeled data [12].

In this work, we created a foundational model for blood parasites based on a SSL approach, enabling the acquisition of visual representations without dependance of labeled images. This foundational model serves as the basis for a novel ‘multiplex’ AI algorithm, specifically designed to concurrently identify diverse parasites within a single blood sample. Our proposed approach offers a robust solution, especially beneficial for classifying microscopy images with limited labeled data.

## 2. Methodology

### 2.1. Dataset

An extensive multi-center clinical dataset of blood samples from patients suspected of parasitic infection (filariae, malaria, chagas disease, and leishmaniasis) was collected and digitized with a smartphone attached to a conventional microscope using a 3D-printed adapter (Spotlab, Spain) [13]. Blood samples from 332 subjects were digitized, yielding 11,956 microscope field-of-view images (FoV). To increase the variability of our dataset, we included a public dataset of Chagas with 675 images [7], thus forming a large database with images from 4 different sites. Our database includes images with different magnifications: 10× (∼0.81 µm/pixel), 40× (∼0.20 µm/pixel), 100× (∼0.081 µm/pixel), each FoV measures ∼2700 x 2700 pixels.

To tailor the dataset for our classification objectives and maintain granularity, we divided each FoV into a structured 3×3 grid layout, resulting in a total of 105,075 cropped patches that were resized to 300x300 pixels. FoV images were labeled by expert microscopists. From those images, we curated a subset of 15,268 labeled patches that contained parasites across 11 unique classes, as detailed in **Table 1**.

**Table 1.** Labels description.

|  |  |
| --- | --- |
| Number of labeled patches | 15,268 |
| Microfilariae (filariae) | 2,882 |
| *L. loa* (filariae) | 3,677 |
| *M. perstans* (filariae) | 982 |
| *W. bancrofti* (filariae) | 546 |
| *B. malayi* (filariae) | 620 |
| *P. vivax* (malaria) | 450 |
| *P. falciparum* (malaria) | 2,297 |
| *P. knowlesi* (malaria) | 762 |
| *P. ovale* (malaria) | 433 |
| *P. malariae* (malaria) | 151 |
| *T. cruzi* (chagas disease) | 2,468 |

These 15,268 images were used for training and assessing model classification performance. The annotated images were further divided into training and validation sets using a 5 fold cross-validation split at patient level and stratified by species distribution. On the other hand, the remaining 89,600 unlabeled patches were used for training the self-supervised phase of the proposed approach. To maintain the integrity of the training process, the labeled dataset for training the classifier and the unlabeled dataset were segregated so that no image patches are used in both the self-supervised phase and the subsequent classification.

### 2.2. Overview of the proposed method

Utilizing large-scale pretrained models has been proven to offer diverse advantages, such as enhancing downstream task performance and minimizing the required labeled data for subsequent training tasks [14], [15]. **Figure 1** illustrates the proposed pipeline. To summarize, we first initialize a backbone encoder network with weights from ImageNet, followed by transferring these weights to train the SSL network. During this phase, the network undergoes training using a large, unlabeled dataset of microscopy blood images, allowing it to autonomously acquire visual representations of blood parasites without the need for manual annotations. Subsequently, transfer learning is once again applied, culminating in the network’s final training for the downstream task using the labeled dataset. The primary objective of this final network is to classify each microscopy image, discerning the presence of the previously defined 11 possible blood parasites. This streamlined pipeline optimizes the advantages of pretrained models and self-supervised learning, ensuring robust performance while minimizing the demand for extensive labeled data.

**Fig. 1.**
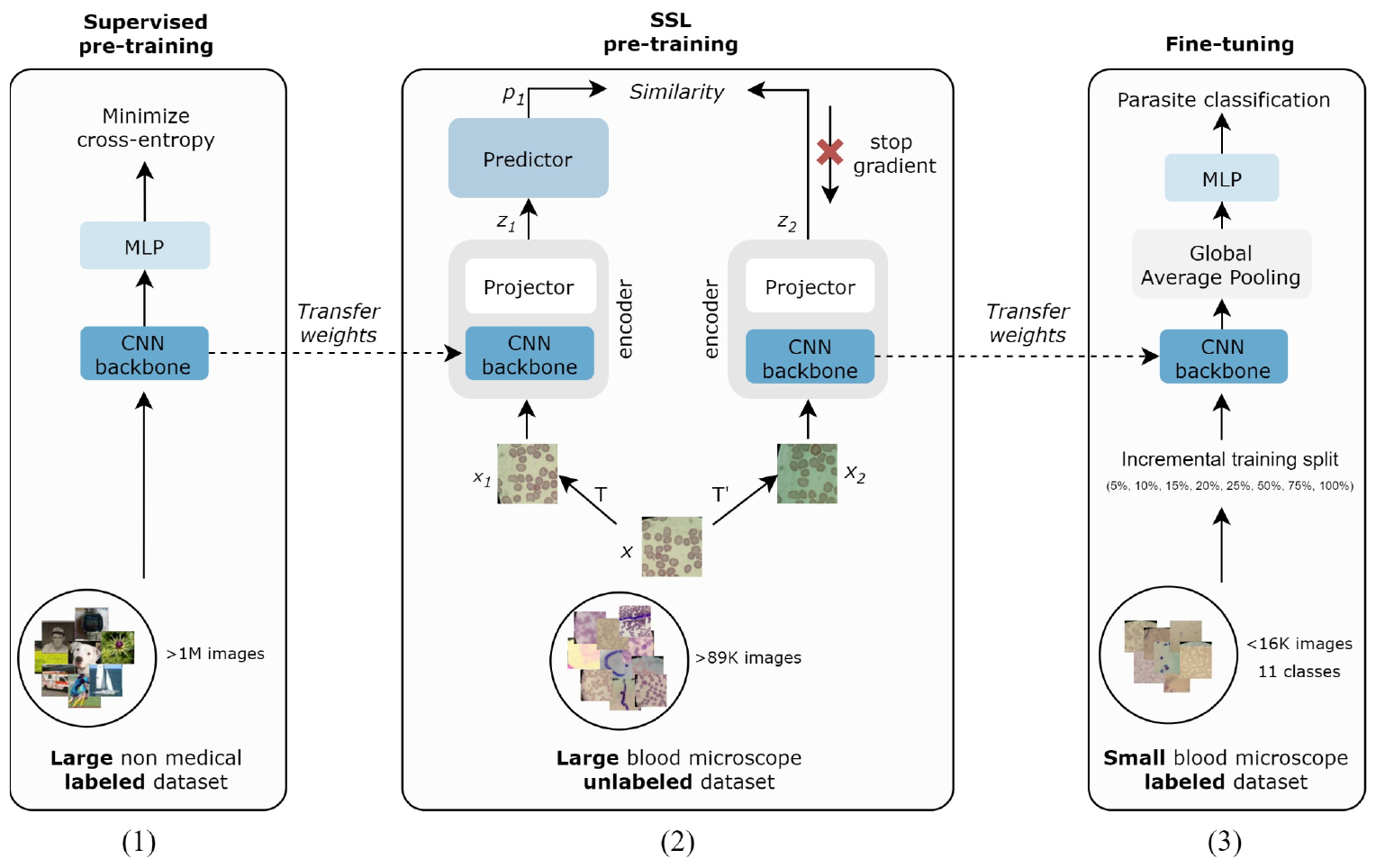
Proposed methodology, comprising: (1) supervised representation learning on a large-scale dataset of labeled natural images such as ImageNet; (2) self-supervised contrastive representation learning on a large unlabelled microscopy dataset; and (3) supervised fine-tuning on manually labeled dataset for downstream task.

### 2.3. Image representation with self-supervised learning

We used SSL for improving blood microscopy image representation by training a feature extractor (backbone, ResNet50) in order to increase data generalization, by leveraging unlabeled datasets. In this work, we implemented a method based on SimSiam [16], a non-contrastive SSL algorithm that, in contrast to SimCLR [17] or MoCO [18], does not need negative samples nor large batches and tries to optimize the similarity among images using only positive pairs. In order to learn meaningful representations in a self-supervised manner, we used a siamese network which tries to maximize the similarity between two different views (transformations) of the same original image. For this purpose, given an image *x*, two different views (*x*_1_, *x*_2_) are generated with random augmentations *T* and *T*’. Such transformations hold the potential to influence the efficacy of a pre-trained model increasing its generalization capability. The transformations selected for our investigation include random cropping, color jittering, and random flipping, all of which align with the augmentation strategies utilized within the SimCLR framework [17] and are commonly adopted as baseline augmentations in a wide array of SSL algorithms. Hyperparameters of these transformations (e.g. crop scale or brightness) were selected with careful consideration of the clinical context of microscopy images, aiming to prevent the generation of non-realistic results as well as to ensure color and space invariance. The generated views are processed by an encoder, formed by a convolutional backbone and a 3-layer multi layer perceptron (MLP) projector to generate image embeddings *z*_1_and *z*_2_. Then, each embedding, *z*, is processed by a 2-layer bottleneck MLP predictor producing embeddings *p*_1_ and *p*_2_. Loss function was defined based on the negative cosine similarity between the vectors *z*_1_ and *p*_2_:

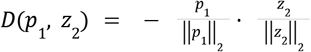

A key factor of this algorithm is the introduction of stop gradients to avoid representation collapse as seen in **Figure 1** By updating only the gradient of the network output taking into account the output of the predictor, it avoids the production of constant value for every image leading to no improvement in the representation learning [19]. This process is done once per each pair of views(*x*_1_, *x*_2_) leading to the following symmetrized loss function:

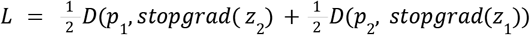

### 2.4. Incremental training for parasite classification

Once the backbone architecture is trained via self-supervised learning, weights are transferred and fine-tuned for the downstream classification task. At this point, we tested two different approaches. The first approach was based on only allowing the training of the weights of the last convolutional layers and those from the classification dense layers, and leaving the weights of the first convolutional layers frozen (*linear probe*). In the second approach, we allow fine-tuning all weights of the architecture.

To assess the impact of self-supervised learning (SSL) in a constrained labeled dataset scenario, we conducted an incremental training experiment, augmenting the training size in steps of 5%, 10%, 15%, 25%, 50%, 75% and 100%.

Employing 5-fold cross-validation, we systematically evaluated parasite classification performance across 11 classes, comparing the SSL pretrained backbone to a baseline ResNet50 classifier with ImageNet weights.

All classification networks were implemented to optimize the cross-entropy loss. During training, various data augmentation techniques were applied such as random flipping, brightness and saturation transformations. To handle the imbalanced distribution across the 11 classes of our labeled dataset and to reduce potential overfitting to the majority classes, we weighted the loss function according to class distributions.

## 3. Experiments and Results

All networks were implemented on Tensorflow (v2.11.0), and trained on a workstation equipped with a NVIDIA A10G 24GB GPU, 32 GB RAM and 8 CPUs.

### 3.1. Self-supervised pre-training

ResNet50 backbone was initialized with ImageNet weights in order to speed up the pre-training. The previously described loss function for training the SSL network was optimized with SGD method with 1e-4 weight decay and 0.9 momentum, with a cosine decayed learning rate of 0.01× (*batch size* / 256). We used a batch size of 32 and pre-trained the model over a span of 25 epochs.

Figure 2. reveals the model’s capability to extract relevant features illustrating a high degree of intra-class similarity and a significant inter-class separation in the feature latent space. This shows that the learned representations are meaningful to distinguish the multiple parasite species.

**Fig. 2.**
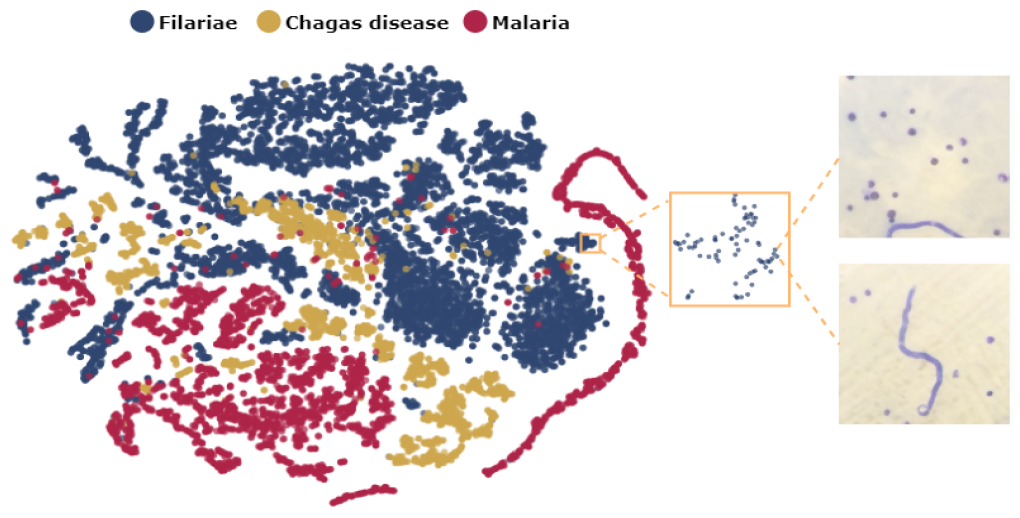
tSNE visualization of the features extracted by SimSiam.

### 3.2. Incremental training and classification results

We trained with the Adam optimizer with 1e-4 weight decay, with an initial learning rate of 0.01 with Cosine Decay. All models were evaluated on the same validation set, and the performance was assessed using the macro (unweighted) F1 score. **Figure 3** shows that leveraging the foundational SSL weights enhances performance compared to training from scratch, particularly in scenarios with limited labeled data. **Table 2** provides a breakdown of model performance, including SSL performance when fine-tuning all weights rather than just the last layers.

**Table 2.** Model performance (macro F1-score) of the proposed model and different weight fine-tuning strategies.

| Method | Supervised | SSL pre-training |  |
| --- | --- | --- | --- |
|  |  | Fine tuning | Linear probe |
| 5% | 0.056 ± 0.056 | 0.195 ± 0.110 | **0.689 ± 0.022** |
| 10% | 0.294 ± 0.133 | 0.518 ± 0.037 | **0.757 ± 0.019** |
| 15% | 0.512 ± 0.138 | 0.547 ± 0.047 | **0.784 ± 0.030** |
| 20% | 0.605 ± 0.114 | 0.671 ± 0.045 | **0.813 ± 0.013** |
| 25% | 0.671 ± 0.045 | 0.698 ± 0.058 | **0.823 ± 0.009** |
| 50% | 0.814 ± 0.026 | 0.827 ± 0.029 | **0.873 ± 0.011** |
| 75% | 0.864 ± 0.020 | 0.882 ± 0.010 | **0.889 ± 0.006** |
| 100% | 0.890 ± 0.008 | 0.892 ± 0.010 | **0.901 ± 0.016** |

**Fig. 3.**
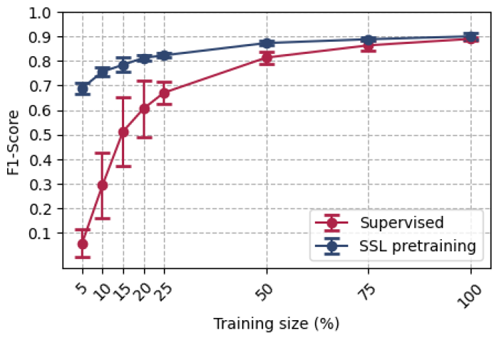
Comparative performance (macro F1-score) analysis illustrating the impact of employing the SSL methodology versus not using it, across an incremental training experiment.

**Table 2** illustrates that freezing initial weights (linear probe) when little labeled data is available significantly boosts performance. Full-model fine-tuning with scarce data risks losing vital pretrained features and may lead to overfitting. In contrast, restricting fine-tuning to the latter layers markedly improves performance, utilizing stable features from self-supervised pre-training for task adaptation. Detailed performance of our proposed method for each class under study is shown in **Figure 4**, where we show that our model with just ∼100 labels can achieve a mean F1 score of 0.798 and a deviation of 0.074 across the 11 parasite species within our dataset.

**Fig. 4.**
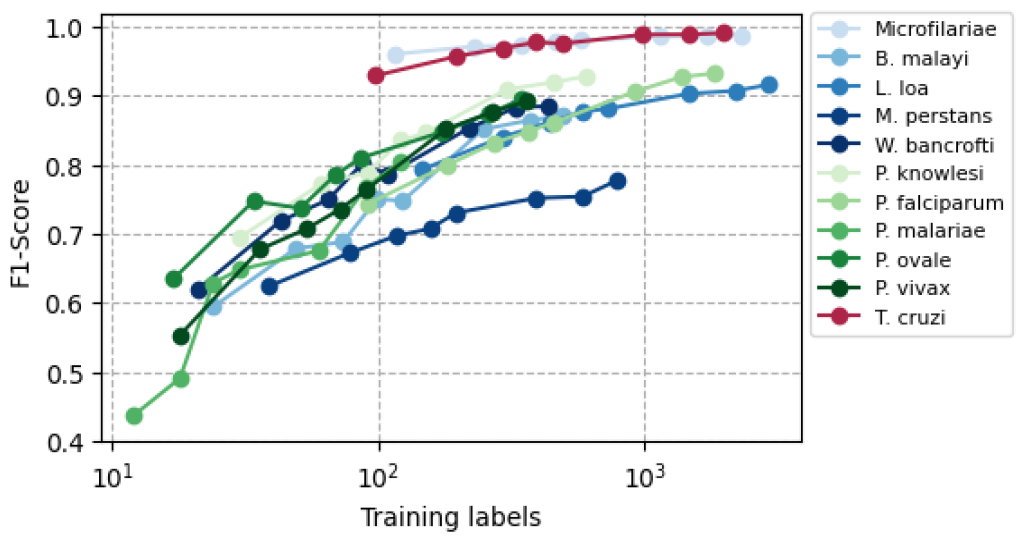
F1-Score comparison for each class using the SSL model.

Figure 5. provides a detailed depiction of the outcomes presented in the previous figure, specifically highlighting our model performance across individual classes when the number of training labels is close to 100.

**Fig. 5.**
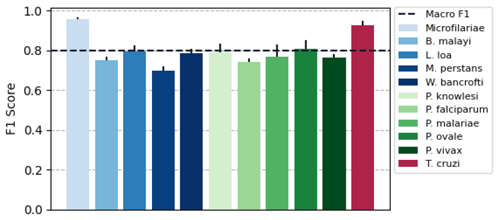
F1-Score when training labels is ∼100 for each class

In order to assess our SSL model’s generalization capability, we tested it on an independent malaria blood sample classification task using a public database of 2,025 images (1,751 for training, 274 for testing) across 7 classes (various malaria species and maturation stages) [20]. Transferring weights from our SSL model improved classification (macro F1-score), from 38.90% (using a ResNet50 initialized with ImageNet weights) to 83.18%.

## 4. Conclusions

We propose a data-efficient methodology for classifying multiple blood parasite species using SSL. This approach leverages large unannotated databases to learn meaningful features, increasing model performance when limited annotated data is available. Our experiments show better performance of our SSL based model over an ImageNet pretrained network. We have found that using a self-supervised methodology with >89000 images, is then enough to have ∼100 labels per parasite class to get an F1 score of ∼0.8, and we expect ∼1000 labels to achieve F1 of ∼0.9. Remarkably, parasite classes exhibit a similar trajectory regarding performance as training data increases. This characterization of the expected performance is very important in real world deployments when designing a clinical AI protocol to include in the diagnostic system new parasites which might have limited data available-either because of the low prevalence or because it is hard to reach the populations suffering from these diseases.

Our work is particularly relevant for the diagnosis of neglected tropical diseases (NTDs), where labeled data is often limited. We diverge from the existing literature focused on specific parasite detection by formulating a generalized AI framework for multiple blood parasite species in a single model, aligning with the real-world need for efficient, all-encompassing diagnostic tools. Our research has significant real-world application potential, consolidating multiple diagnostic models into a single one, which can be deployed in a lightweight mobile application optimizing both storage and computational requirements.

While promising, our current patch-based method overlooks factors such as parasite abundance and distribution within the field of view, meriting future inquiry. Nonetheless, this research marks a step toward achieving the World Health Organization’s performance benchmarks for in-vitro diagnostic devices, leveraging AI to combat parasitic diseases.

## 5. Compliance with ethical standards

Ethical approval was obtained from the Research Ethics Committee (REC) Instituto de Salud Carlos III, Spain (CEI PI 74_2020) and the favorable report of the bioethics committee of the faculty of medicine of the Universidad Mayor de San Simón (Cochabamba, Bolivia).

## Data Availability

Images and labels can be shared for research purposes upon request. Please contact Miguel Luengo-Oroz.

## 6. Acknowledgements

We extend our sincere gratitude to Jose Miguel Rubio and Maria Flores Chavez and their colleagues from Instituto de Salud Carlos III and Mary Cruz Torrico and colleagues from Universidad Mayor de San Simón for their invaluable contribution to this research by providing essential data. We would also like to thank Mauro César Cafundó Morais and colleagues for publishing the Chagas dataset.

This project has been partially funded by the European Union’s Horizon 2020 research and innovation programme (grant agreement No 881062) and the Bill and Melinda Gates Foundation (grant number Edge-Spot project INV-051355). LL was supported by a predoctoral grant IND2019/TIC-17167 (Comunidad de Madrid).

## References

[1] S. Naicker, J. Plange-Rhule, R. C. Tutt, and J. B. Eastwood, “Shortage of healthcare workers in developing countries--Africa.,” Ethn. Dis., vol. 19, no. 1 Suppl 1, pp. S1–60, 2009.

[2] Global strategy on human resources for health: Workforce 2030. 2020.

[3] D. Das et al., “Field evaluation of the diagnostic performance of EasyScan GO: a digital malaria microscopy device based on machine-learning.,” Malar. J., vol. 21, no. 1, p. 122, Apr. 2022, doi: 10.1186/s12936-022-04146-1.

[4] H. Yu et al., “Patient-level performance evaluation of a smartphone-based malaria diagnostic application.,” Malar. J., vol. 22, no. 1, p. 33, Jan. 2023, doi: 10.1186/s12936-023-04446-0.

[5] J. Pfeil, A. Nechyporenko, M. Frohme, F. T. Hufert, and K. Schulze, “Examination of blood samples using deep learning and mobile microscopy.,” BMC Bioinformatics, vol. 23, no. 1, p. 65, Feb. 2022, doi: 10.1186/s12859-022-04602-4.

[6] M. S. Davidson et al., “Automated detection and staging of malaria parasites from cytological smears using convolutional neural networks.,” Biol. Imaging, vol. 1, p. e2, Aug. 2021, doi: 10.1017/S2633903X21000015.

[7] M. C. C. Morais et al., “Automatic detection of the parasite Trypanosoma cruzi in blood smears using a machine learning approach applied to mobile phone images.,” PeerJ, vol. 10, p. e13470, May 2022, doi: 10.7717/peerj.13470.

[8] N. Sanchez-Patino, A. Toriz-Vazquez, N. Hevia-Montiel, and J. Perez-Gonzalez, “Convolutional neural networks for chagas’ parasite detection in histopathological images.,” Annu. Int. Conf. IEEE Eng. Med. Biol. Soc., vol. 2021, pp. 2732–2735, Nov 2021, doi:10.1109/EMBC46164.2021.9629563.

[9] C. Gonçalves et al., “Detection of Human Visceral Leishmaniasis Parasites in Microscopy Images from Bone Marrow Parasitological Examination,” Appl. Sci., vol. 13, no. 14, p. 8076, Jul. 2023, doi: 10.3390/app13148076.

[10] U. Ornellas-Garcia, P. Cuervo, and F. L. Ribeiro-Gomes, “Malaria and leishmaniasis: Updates on co-infection.,” Front. Immunol., vol. 14, p. 1122411, Feb. 2023, doi: 10.3389/fimmu.2023.1122411.

[11] P. Wilairatana, K. U. Kotepui, W. Mala, K. Wangdi, and M. Kotepui, “Prevalence, probability, and characteristics of malaria and filariasis co-infections: A systematic review and meta-analysis.,” PLoS Negl. Trop. Dis., vol. 16, no. 10, p. e0010857, Oct. 2022, doi: 10.1371/journal.pntd.0010857.

[12] S.-C. Huang, A. Pareek, M. Jensen, M. P. Lungren, S. Yeung, and A. S. Chaudhari, “Self-supervised learning for medical image classification: a systematic review and implementation guidelines.,” npj Digital Med., vol. 6, no. 1, p. 74, Apr. 2023, doi: 10.1038/s41746-023-00811-0.

[13] E. Dacal et al., “Mobile microscopy and telemedicine platform assisted by deep learning for the quantification of Trichuris trichiura infection.,” PLoS Negl. Trop. Dis., vol. 15, no. 9, p. e0009677, Sep. 2021, doi: 10.1371/journal.pntd.0009677.

[14] M. Nielsen, L. Wenderoth, T. Sentker, and R. Werner, “Self-Supervision for Medical Image Classification: State-of-the-Art Performance with ∼100 Labeled Training Samples per Class.,” Bioengineering (Basel), vol. 10, no. 8, Jul. 2023, doi: 10.3390/bioengineering10080895.

[15] S. Azizi et al., “Robust and Efficient Medical Imaging with Self-Supervision,” arXiv, 2022, doi: 10.48550/arxiv.2205.09723.

[16] X. Chen and K. He, “Exploring Simple Siamese Representation Learning,” arXiv, 2020, doi: 10.48550/arxiv.2011.10566.

[17] T. Chen, S. Kornblith, M. Norouzi, and G. Hinton, “A Simple Framework for Contrastive Learning of Visual Representations,” arXiv, 2020, doi: 10.48550/arxiv.2002.05709.

[18] K. He, H. Fan, Y. Wu, S. Xie, and R. Girshick, “Momentum Contrast for Unsupervised Visual Representation Learning,” arXiv, 2019, doi: 10.48550/arxiv.1911.05722.

[19] C. Zhang, K. Zhang, C. Zhang, T. X. Pham, C. D. Yoo, and S. Kweon, “How Does SimSiam Avoid Collapse Without Negative Samples? A Unified Understanding with Self-supervised Contrastive Learning,” arXiv, 2022, doi: 10.48550/arxiv.2203.16262.

[20] MalariaSystem, “malaria diagmal Dataset.” https://universe.roboflow.com/malariasystem/malaria_diagmal (accessed Nov. 22, 2023).

